# Association between women’s empowerment and demand for family planning satisfied among Christians and Muslims in multi-religious African countries

**DOI:** 10.1101/2023.08.06.23293712

**Authors:** Franciele Hellwig, Yohannes Dibaba Wado, Aluísio JD Barros

## Abstract

**Background:** Although the levels of demand for family planning satisfied (DFPS) have increased in many countries, cultural norms remain a significant barrier in low- and middle-income countries. In the context of multi-religious African countries, our objective was to investigate intersectional inequalities in DFPS by modern or traditional contraceptives according to religion and women’s empowerment.

**Methods:** Analyses were based on Demographic and Health Surveys carried out between 2010 and 2021 in African countries. Countries with at least 10% of Muslims and Christians were selected to analyze inequalities in family planning. The religious groups were characterized by wealth, area of residence, women’s age, and women’s empowerment. The mean level of empowerment was estimated for each religious group, and multilevel Poisson regression was used to assess whether demand for family planning satisfied varied based on the level of women’s empowerment among Muslims and Christians.

**Results:** Our study sample of 14 countries comprised 35% of Muslim and 61% of Christian women. Christians had higher levels of empowerment across all three domains compared to Muslims and women with no/other religion. DFPS was also higher among Christians (57%) than among Muslims (36%). Pooled analysis indicated a consistent association between DFPS and women’s empowerment, with higher prevalence ratios among Muslims than Christians, especially in the decision-making domain.

**Conclusions:** The gap between Muslims and Christians in DFPS significantly reduced as the level of empowerment increased. It highlights the importance of understanding and addressing cultural factors sensibly and respectfully to satisfy the demand for family planning services.

**Research in context:** *What is already known on this topic:*

– Prior studies have demonstrated that there is a complex and variable relationship between religion and family planning beliefs in sub-Saharan Africa.
– Higher fertility and lower use of contraceptives among Muslims than among Christians were documented in several African countries.
– The literature on the relationship between women’s empowerment and contraceptive use is mixed, with some studies identifying null associations and others finding positive associations.
– A qualitative study conducted in Tanzania has identified that both religion and gender dynamics influence family planning practices.

*What this study adds:*

– Our study builds on the prior literature by using data from 148,989 women to investigate intersectional inequalities in demand for family planning satisfied by religion and women’s empowerment across multiple countries.
– While Muslim was the group with lower levels of women’s empowerment in the three SWPER domains, African Christians were on average more empowered than the average of women from all low- and middle-income countries.
– Lower levels of DFPS were also identified among Muslim women, however, the coverage increased significantly with their empowerment. With the highly empowered Muslims having higher mDFPS than the highly empowered Christians.
– Among the SWPER domains, stronger effects were found in the decision-making domain.
– Higher pooled prevalence ratios were identified when considering traditional methods instead of modern contraceptives.

*How this study might affect research, practice, or policy:*

– This study can help researchers, policymakers, and policy managers to better understand how social norms affect the use of family planning services. While religious beliefs can be a challenge to family planning, promoting gender equity and empowering women can help to mitigate these challenges and improve access to reproductive healthcare services for women.

## Introduction

Although considerable strides have been made to increase levels of the demand for family planning satisfied (DFPS) with modern methods in low- and middle-income countries (1, 2), there are still persistently low levels of coverage in certain countries and among certain groups of women, such as the poorest, youngest age groups, Muslims, and those with lower levels of education (3). Increasing family planning use is especially challenging in sub-Saharan Africa, where a significant portion of the population lives in extreme poverty, and health services fail to reach many women (4–6). Nevertheless, the influence of cultural factors such as gender inequality, early marriage, fertility expectations, and negative attitudes toward family planning practices is progressively assuming a central role (3, 7–9).

Gender inequality is crucial in shaping women’s access to and utilization of reproductive health services. This is influenced by various factors, including women’s limited participation in household decisions, restricted decision-making capabilities related to their fertility, lack of mobility, financial constraints, and the occurrence of domestic violence (7, 10, 11). While evidence on the impact of women’s empowerment on family planning utilization remains inconclusive due to methodological limitations, studies generally suggest that higher levels of empowerment, particularly decision-making and freedom of movement, are associated with increased family planning utilization (12–15).

The relationship between family planning and women’s empowerment is often intertwined with societal and personal values (16), religion being one of them. Evidence suggests religious leaders can influence individual behavior, including their reproductive health choices (8, 17). Condemnation of modern contraception due to religious beliefs has been linked to lower family planning use (17–20). However, religions are not similar in their views on contraceptive usage, and they can affect practices differently (17, 21). Also, religious teachings affect individual choices differently across affiliations and social contexts. Some studies showed that Christian women tend to present higher family planning utilization than Muslims (3, 22).

The relationship between women’s empowerment, religion, and family planning utilization is a complex subject that has not been sufficiently explored in the literature. This study explores the independent effects of women’s empowerment and religion on demand for family planning satisfied and whether the intersection of these effects is relevant. The study used data from African countries where Muslim and Christian women are found in large enough proportions and focused on family planning with modern and traditional contraceptives.

## Materials and methods

### Data sources

We used publicly available data from the most recent Demographic and Health Surveys (DHS) carried out from 2010 to 2021. In the 14 African countries with available data on religion, women’s empowerment, and demand for family planning satisfied, women were classified into Muslim, Christian, or other (other faiths or unaffiliated) according to their report. The analyses included only those countries where the proportion of Christian women and Muslim women individually constituted at least 10% of the total population. The analyses were based on currently partnered women aged 15-49.

### Outcome

Our main outcome was demand for family planning satisfied (DFPS), defined as the proportion of women in need of contraception who were using (or whose partner was using) a contraceptive method. A woman was considered in need of contraception if she was fecund and did not want to become pregnant within the next two years or was unsure about whether or when she wanted to become pregnant. Pregnant women with a mistimed or unintended pregnancy were also considered in need of contraception.

DFPS was explored according to the type of contraceptive method used, modern or traditional. Modern contraceptives were defined as medical procedures or technological products (23), including oral contraceptive pills, injections, male and female condoms, diaphragms, spermicidal agents, emergency contraception, intrauterine devices (IUD), implants, and sterilization (female or male). Traditional methods included all calendar-based methods, lactational amenorrhea, and withdrawal.

### Stratifiers and descriptive variables

Women’s empowerment was measured using the SWPER Global (24, 25), an individual-level indicator estimated for women aged 15-49 who are married or in a union. The SWPER was derived using principal component analyses based on 14 DHS questions covering three domains of women’s empowerment: (I) social independence, related to access to information, education, and age of marriage and first birth; (II) decision-making, related to making decisions on important household matters; and (III) attitude towards violence, related to how much the woman rejects domestic violence against the wife. The resulting scores were standardized so that positive values represent above-average levels of empowerment across LMICs used to derive the indicator, while negative values represent the opposite. The zero value represents average empowerment. Based on the continuous SWPER scores, each domain was categorized into low, medium, and high levels of empowerment based on approximate tertiles.

### Statistical analyses

We conducted pooled individual-level analyses across the selected countries using multilevel Poisson regression to evaluate the overall association between religion, women’s empowerment, and DFPS by traditional and modern methods. The multilevel model was structured in two levels: women at level one and countries at level two. The second level was included only to consider the within-country correlation, the model did not include contextual factors in the adjustment. The interaction between women’s empowerment and religion was investigated in the regression models and the differences in the association between Christian and Muslim women were assessed using separate religion-specific regression models.

All analyses were performed using Stata software version 18.0 (StataCorp LLC, College Station, TX) and adjusted for the sample design, including sample weights, clusters, and strata. All analyses relied on publicly available anonymized databases. Institutions and national agencies in each country obtained ethics approval for the surveys.

## Results

### Description of the selected countries

A total of 14 countries that fulfilled the inclusion criteria were included in the analysis. Ten countries were from West and Central Africa, and four were from Eastern and Southern Africa. Christians were the majority in eight countries (Malawi, Uganda, Liberia, Ghana, Togo, Mozambique, Benin, and Ethiopia), while Muslims were the majority in five (Guinea, Sierra Leone, Burkina Faso, Nigeria, and Chad). In Côte d’Ivoire, Christians represented 46% of the population, while Muslims accounted for 40%. The other or unaffiliated religious groups were generally small, except in Togo (26%), Mozambique (21%), Benin (16%), and Côte d’Ivoire (16%) (Table 1).

**Table 1.** Distribution of religious groups and demand for family planning satisfied in the 14 study countries.

| Country | Religion |  |  | Demand for family planning satisfied (%) |  |  | N (unweighted) |
| --- | --- | --- | --- | --- | --- | --- | --- |
|  | Muslim (%) | Christian (%) | Other/<br>unaffiliated (%) | Traditional | Modern | Any |  |
| Benin (2017) | 27.8 | 55.9 | 16.3 | 6.7 | 24.9 | 31.6 | 11170 |
| Burkina Faso (2010) | 61.7 | 31.8 | 6.5 | 3.0 | 36.6 | 39.6 | 13392 |
| Chad (2014) | 45.8 | 51.1 | 3.1 | 5.4 | 14.0 | 19.4 | 13439 |
| Côte d'Ivoire (2011) | 40.9 | 43.4 | 15.7 | 10.8 | 25.6 | 36.4 | 6453 |
| Ethiopia (2016) | 30.3 | 68.4 | 1.3 | 1.4 | 60.2 | 61.6 | 9824 |
| Ghana (2014) | 14.6 | 79.9 | 5.4 | 7.3 | 38.4 | 45.6 | 5456 |
| Guinea (2018) | 84.8 | 12.2 | 3.0 | 11.4 | 19.7 | 31.1 | 7812 |
| Liberia (2019) | 12.7 | 85.6 | 1.7 | 1.5 | 40.5 | 42.0 | 4654 |
| Malawi (2015) | 11.6 | 87.8 | 0.7 | 1.5 | 74.4 | 75.9 | 15952 |
| Mozambique (2015) | 18.7 | 60.4 | 20.9 | 4.9 | 48.0 | 52.9 | 4363 |
| Nigeria (2018) | 45.9 | 53.7 | 0.4 | 14.6 | 30.4 | 45.0 | 28888 |
| Sierra Leone (2019) | 77.4 | 22.6 | 0.0 | 0.9 | 44.9 | 45.8 | 9837 |
| Togo (2013) | 16.1 | 58.0 | 25.9 | 4.3 | 32.2 | 36.5 | 6360 |
| Uganda (2016) | 13.7 | 85.3 | 0.9 | 7.5 | 49.7 | 57.1 | 11379 |

DFPS ranged from 19% in Chad to 76% in Malawi, with the modern method being the most commonly used in all countries. DFPS with traditional methods was higher than 10% only in Nigeria (15%), Guinea (11%), and Côte d’Ivoire (11%) (Table 1).

Table 2 describes the religious groups according to key characteristics. No meaningful difference in terms of women’s age was observed between the three religious groups. Muslims and Christians exhibited comparable distributions in terms of area of residence and wealth quintiles. Women with other religions or unaffiliated were mostly rural (76%); and poorer (60% were from the two poorest wealth quintiles). Regarding women’s empowerment, Christians presented higher levels of empowerment across all three SWPER domains, especially in the decision-making domain, where 55% of Christians were in the high level of empowerment while only 36% of the Muslims were highly empowered.

**Table 2.** Description of the main religious groups in terms of wealth, area of residence, women’s age, and women’s empowerment.

|  | Religion |  |  | p value* |
| --- | --- | --- | --- | --- |
|  | Muslims | Christians | Other/<br>unaffiliated |  |
| **Area of residence** |  |  |  | <0.001 |
| Urban | 39.2 | 35.1 | 24.3 |  |
| Rural | 60.8 | 64.9 | 75.7 |  |
| **Wealth** |  |  |  | <0.001 |
| Poorest | 20.1 | 17.6 | 35.6 |  |
| Poorer | 18.4 | 19.7 | 23.9 |  |
| Middle | 19.2 | 20.2 | 18.3 |  |
| Wealthier | 20.6 | 20.3 | 13.0 |  |
| Wealthiest | 21.7 | 22.2 | 9.1 |  |
| **Women's age** |  |  |  | <0.001 |
| 15-19 yrs | 5.0 | 4.6 | 5.3 |  |
| 20-34 yrs | 57.1 | 58.7 | 56.3 |  |
| 35-49 yrs | 37.9 | 36.6 | 38.4 |  |
| **Women's empowerment** |  |  |  |  |
| Social independence |  |  |  | <0.001 |
| Low | 44.3 | 30.4 | 47.9 |  |
| Middle | 30.6 | 39.3 | 34.8 |  |
| High | 25.1 | 30.3 | 17.3 |  |
| Attitude to violence |  |  |  | <0.001 |
| Low | 28.1 | 21.5 | 26.8 |  |
| Middle | 18.3 | 19.0 | 21.3 |  |
| High | 53.6 | 59.5 | 51.8 |  |
| Decision-making |  |  |  | <0.001 |
| Low | 32.4 | 13.0 | 23.0 |  |
| Middle | 32.0 | 31.7 | 38.1 |  |
| High | 35.5 | 55.4 | 38.9 |  |

\*Pearson chi-squared

The mean level of empowerment according to each SWPER domain and religion is presented in Figure 1. Muslims had lower levels of empowerment than Christians in all three domains, with negative values in all the SWPER domains, indicating a level of empowerment below the global average. Christians had higher levels of empowerment than average in the decision-making domain and levels close to the average in attitude to violence and social independence.

**Figure 1.**
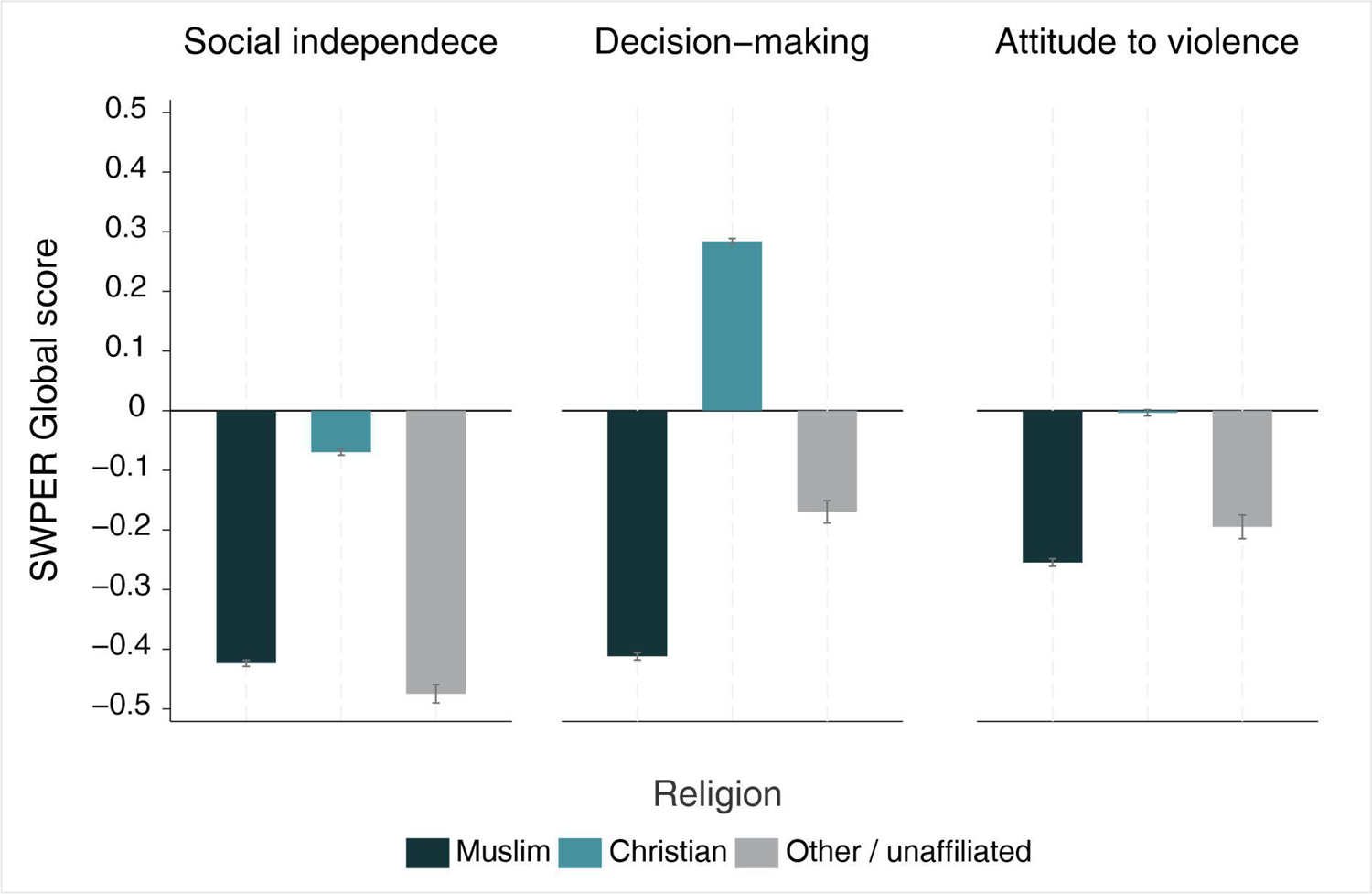
Mean level of women’s empowerment according to religion.

### DFPS according to women’s empowerment and religion

Figure 2 presents mDFPS for each SWPER domain and according to each religious group. There are marked differences according to religion, particularly among women with lower levels of empowerment. The largest gaps were identified in the domains of social independence and attitude to violence, where mDFPS was 11 percentage points higher among low empowered Christians in comparison with low empowered Muslims. The pattern was reversed among highly empowered women, where Muslims had higher mDFPS compared to Christians. The largest difference was in social independence, with high empowered Muslims having mDFPS 5 percentage points higher than the high empowered Christians. Irrespective of the level of empowerment and SWPER domain, women with other or no religion had the lowest mDFPS. Smaller disparities were observed when considering the use of traditional contraceptive methods. Results can be found in the supplementary material.

**Figure 2.**
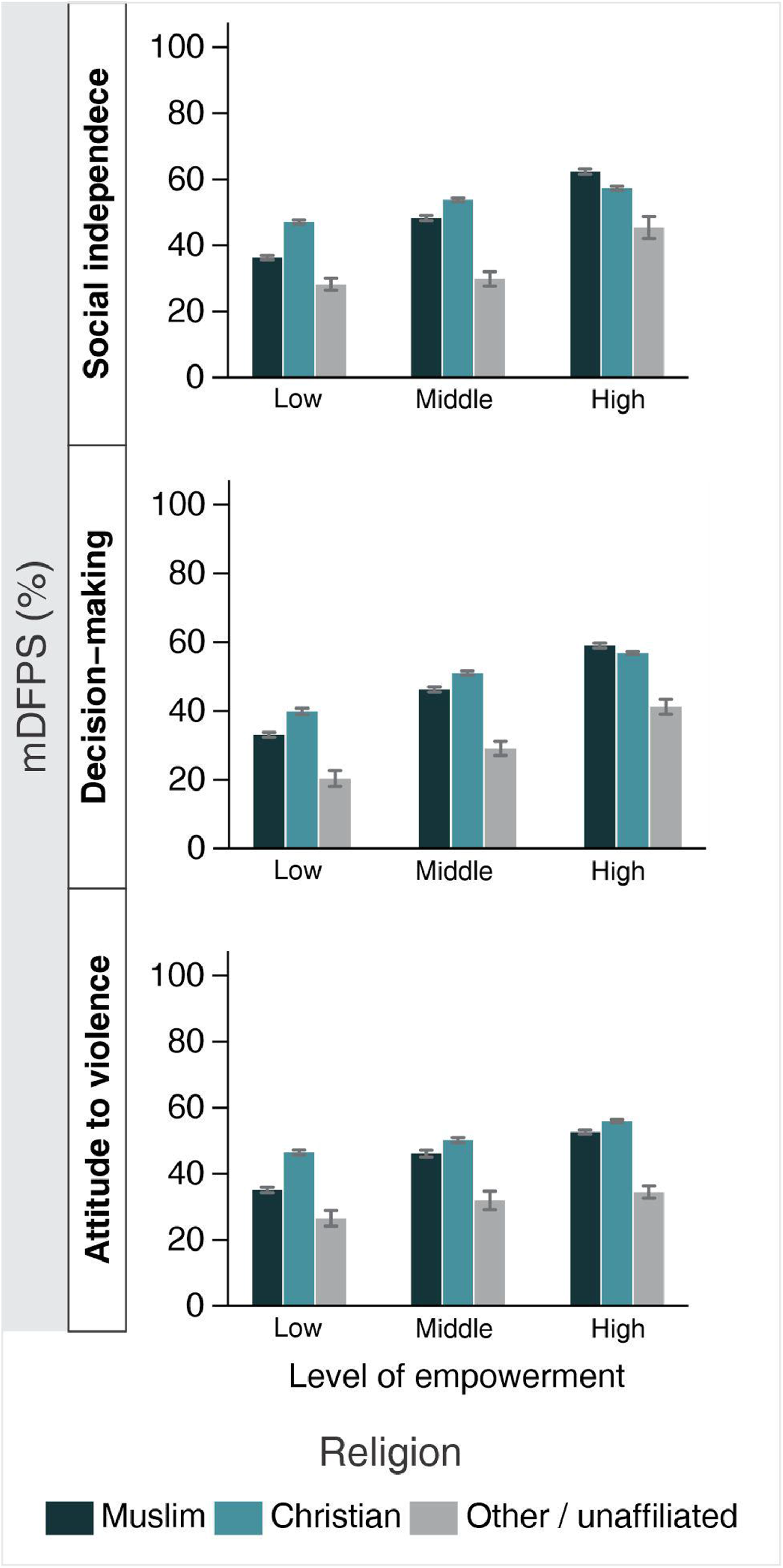
Demand for family planning satisfied by modern methods according to religion and women’s empowerment.

### DFPS and the intersection between religion and women’s empowerment

Figure 3 shows the prevalence ratios comparing high and middle levels of empowerment with low level in terms of DFPS, for Muslims and Christians separately. We found a consistent association between DFPS and religion, with larger prevalence ratios among Muslim women. The prevalence ratios, along with their 95% confidence intervals and interaction p values, are presented in Supplementary Table 1.

**Figure 3.**
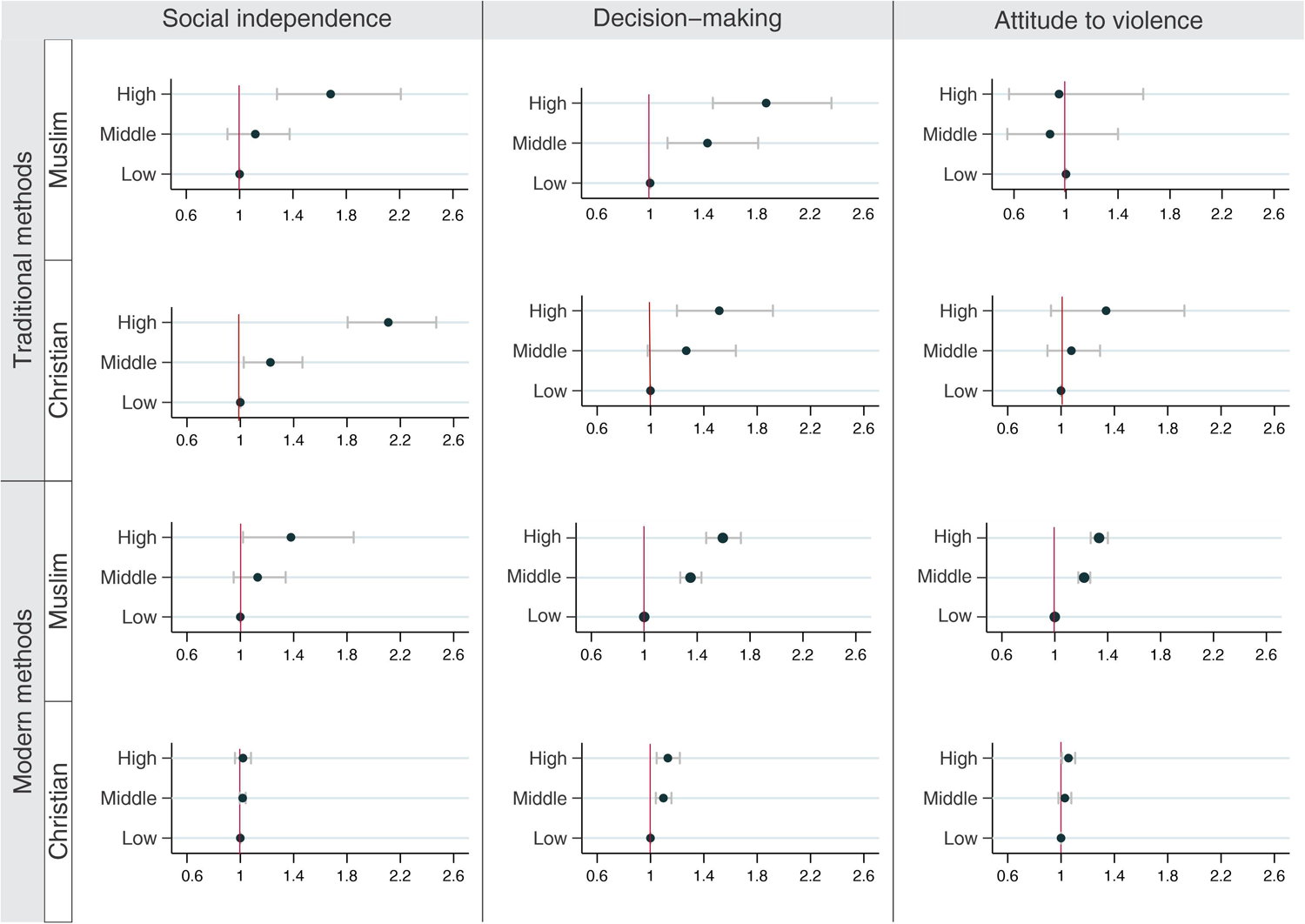
Pooled prevalence ratios for demand for family planning satisfied according to type of method, religion, and level of women’s empowerment., Note: all the interection tests had p<0.001.

We identified a significant relationship between women’s empowerment and DFPS in terms of decision-making power. Regardless of religion or type of contraceptive used, we found that high levels of empowerment were associated with higher DFPS. The highest prevalence ratio was observed among highly empowered Muslims, with their tDFPS being 1.9 times higher than that of the low empowered women. When modern contraceptive methods were considered, the prevalence ratios in this domain decreased significantly among Christians. Among highly empowered Christians, the tDFPS was 50% higher than among the low empowered group, while mDFPS was only 10%. In contrast, the prevalence ratios remained high among Muslims. The mDFPS among highly empowered Muslims was 50% higher compared to the low empowered women.

In the domain of social independence, larger prevalence ratios were observed when considering the use of traditional methods, with Christians exhibiting the highest prevalence ratios. High empowerment among Christians and Muslims was associated with tDFPS 2.1 and 1.7 times higher in comparison with low empowerment. When modern methods were considered, no difference in terms of empowerment level was observed among Christians. Among Muslims, mDFPS was 13% and 38% higher among Muslims in the middle and high levels of empowerment, respectively, than among low empowered Muslims.

No difference was found in tDFPS in the attitude to violence domain between levels of empowerment nor between Muslims and Christians. Among Christians, we also observed no difference in mDFPS across different levels of empowerment. Nevertheless, a significant association between mDFPS and women’s empowerment in terms of attitude towards violence was identified among Muslims. Muslim women in the middle and high levels of empowerment presented levels of mDFPS 20% and 29% higher, respectively, than those of the low empowered group.

## Discussion

In this multi-country study, we found significant intersectional inequalities in DFPS regarding religion and women’s empowerment. African Christians were more empowered than women from other religious groups in Africa and even more empowered than the average women in low- and middle-income countries. Additionally, levels of DFPS were higher among Christians than among other religious groups. The relationship between women’s empowerment and DFPS was stronger among Muslims than Christians, with highly empowered Muslims having higher levels of mDFPS than highly empowered Christians.

Our findings align with previous studies that have shown higher use of contraceptives among Christians compared to Muslims (26–28). Islam allows for different interpretations of the Quran and the Sunnah, leading to variations in the acceptance of contraception (18, 29, 30). While some conservative Muslims argue against any form of contraception, other schools permit fertility regulation for birth spacing (18, 30). Previous studies investigating family planning barriers among African Muslims have identified that due to Islamic beliefs mandating women to grant their husbands unrestricted access to their bodies and the potential resulting pregnancies (19, 20), their concerns about family planning practices goes beyond fate for the afterlife, as they also fear domestic violence if their husbands suspect them of seeking contraceptives (20). Additionally, the perspectives of Muslims on family planning are also shaped by political contexts (8, 20, 31). While people from different religious backgrounds may also question the relevance and origins of birth control ideas, Muslims, in general, tend to be more reluctant to embrace contraception due to their belief that international family planning efforts are politically motivated (8, 31). In agreement, increases in the use of contraception have been documented in countries where family planning policies were implemented without any political connotation (31).

Christian attitudes toward family planning vary significantly depending on the denomination. The three major denominations of Christianity are Roman Catholicism, Eastern Orthodoxy, and Protestantism. Catholicism sees marital sex as a means of procreation and marital unit. These two aspects are seen as inextricably linked. Thus, refusing one will inevitably undermine the other (32). Within Catholicism, periodic abstinence methods are allowed for birth spacing in specific cases, such as when a pregnancy would be associated with health or socioeconomic risks (18, 21, 33). Analogous to Catholicism, Eastern Orthodoxy also sees marital sex as a mean of procreation and marital unit but it differentiates between types of contraceptives, accepting condoms and withdrawal but rejecting hormonal contraceptives and sterilization (18, 21). While some more conservative Protestant denominations are against contraception, Protestantism, in general, takes a more flexible approach to secular issues, including an acceptance of modern contraceptive use (18, 21).

The impact of religion on family planning is influenced by the level of flexibility exhibited by religious representatives and women in navigating religious dogmas. The varying stances taken by representatives of different religions on contraception have been argued to reflect their respective abilities and willingness to preserve their positions in society (31). Evidence suggests that women can reconcile their secular views with religious beliefs, while religious institutions may serve as valuable sources of social power and may offer a platform for discussions on gender roles and healthcare (33, 34). In line with this, our findings indicate that Christian African women were not only more empowered than African women following other religions or unaffiliated, as their level of empowerment in the decision-making domain surpasses the average level observed among women across low- and middle-income countries from all regions. This is a notable finding given the generally poorer socioeconomic conditions in most of the African countries.

The literature consistently underscores the significance of engaging religious leaders in promoting family planning. Consequently, there has been a shift among religious leaders towards increased openness in endorsing and advocating for family planning initiatives, especially by sharing messages about birth spacing through the use of traditional contraceptive methods (35–37). However, it is essential to recognize that the impact of religion on contraceptive use is influenced by a complex interplay of socioeconomic and cultural factors (28, 38). Factors such as education and gender equality not only affect women’s access to and utilization of modern contraceptives but also their ability to properly use traditional contraceptives, which require knowledge of the ovulatory cycle and negotiation within intimate relationships (12–14, 39, 40).

By examining the intersectional inequalities in the use of traditional and modern methods based on three domains of women’s empowerment, we found larger gaps between low- and high-empowered women in demand satisfied by traditional methods in the social independence domain. Variables within this domain, such as women’s education, age at first birth and first cohabitation, and spousal differences in age and education, have been associated with contraceptive use (3, 41–43). When examining the utilization of modern contraceptives, differences were found across all domains, with the decision-making domain yielding more significant results than the other domains. The SWPER decision-making domain includes the woman’s freedom to decide on visiting friends or relatives, on significant household purchases, and on her own healthcare. The influence of women’s decision-making power on contraceptive use has been previously explored, with joint decision-making being associated with higher contraceptive usage (44–47).

Our findings indicate that the effects of women’s empowerment on DFPS were significantly higher among Muslims, with highly empowered Muslims showing the highest levels of DFPS. Studies have consistently found a larger educational gender gap among Muslims compared to followers of other religions (48). Furthermore, countries with more conservative Muslim societies often display less openness to the impacts of modernization (48). Highly empowered Muslims likely represent a specific group characterized by exceptional education levels and a greater willingness to embrace new ideas and technologies.

The women’s empowerment index utilized in this study demonstrated high cross-country consistency, capturing meaningful variations in women’s empowerment levels across diverse cultural and socioeconomic contexts and enhancing the robustness of our findings (24, 25). Additionally, it incorporates several factors strongly associated with family planning coverage, including wealth, area of residence, and women’s education. Acknowledging this, we opted to not adjust our analyses for these relevant variables.

It is important to acknowledge the limitations of this study. One such limitation is the lack of consideration for the heterogeneity within Muslims and Christians, as well as their varying degrees of adherence to religious dogmas. The unavailability of such detailed information in national health surveys limits the depth of our assessment.

Despite these limitations, this study provides valuable insights into the complex relationship between religion, women’s empowerment, and the demand for family planning satisfied. To fully comprehend the influence of religion on family planning, it is essential to consider the cultural, historical, political, and sociodemographic contexts within which it operates. These contextual factors shape how religious groups position themselves in relation to cultural and technological advancements. Moreover, it is important to recognize that religion offers individuals a broad and transcendental perspective to navigate life, while public policies and strategies often adopt a narrow perspective based on their specific objectives. The challenge of developing effective evidence-based health policies lies in striking a balance between the need for a broader perspective, considering these diverse contextual factors, and the inherent limitations imposed by technical feasibility and the availability of financial and non-financial resources for implementation. Investing in studies that delve into these intricate relationships and involving stakeholders in all stages of the policy development process can lead to a more nuanced comprehension of the sociocultural dynamics and facilitate the development of more impactful policies.

## Conclusion

This study demonstrated a significant association between women’s empowerment and DFPS among Muslim women. It also showed that the disparity between Muslims and Christians in DFPS significantly reduced as the level of empowerment increased. By presenting a comprehensive portrayal of this intersectional inequality, our findings could enhance comprehension of the interplay between women’s empowerment, religion, and family planning. It emphasizes the critical need to approach cultural factors with sensitivity and respect to effectively meet the demand for family planning services. Policymakers and healthcare providers should acknowledge the potential impacts of gender roles and religion on a couple’s decision to use contraception and be knowledgeable about the diverse range of family planning options available. By doing so, they can effectively navigate and respond to these cultural and social circumstances in the most optimal manner, ensuring that individuals’ reproductive health needs are met with sensitivity and respectability.

## Contributors

Conceptualization, F.H. and A.J.D.B.; data curation, F.H and A.J.D.B.; formal analysis, F.H.; funding acquisition, A.J.D.B.; investigation, F.H.; methodology, F.H. and A.J.D.B.; project administration, F.H. and A.J.D.B.; supervision, A.J.D.B.; validation, F.H.; visualization, F.H. and A.J.D.B.; writing— original draft, F.H.; writing—review and editing, F.H., Y.D.W., and A.J.D.B. All authors have read and agreed to the published version of the manuscript.

## Declaration of interests

No conflicts of interest were declared by the other authors.

## Patient and public involvement

Patients were not involved.

## Data sharing statement

All data relevant to the study are included in the article or available as supplementary information. The data used in the analyses is publicly available, anonymized and geographically scrambled to ensure confidentiality. More information on DHS can be found at https://dhsprogram.com/, where survey datasets can be obtained.

## Funding

This work was funded by grants from the Bill & Melinda Gates Foundation and ABRASCO (Associação Brasileira de Saúde Coletiva).

## Role of the funding sources

The funders of the study had no role in study design; in the collection, analysis, and interpretation of data; in the writing of the report; and in the decision to submit the paper for publication.

## Data Availability

All data produced in the present work are contained in the manuscript.

## Notes

### Competing Interest Statement

The authors have declared no competing interest.

### Funding Statement

This work was funded by grants from the Bill and Melinda Gates Foundation and ABRASCO (Associacao Brasileira de Saude Coletiva).

### Author Declarations

This paper works specifically with Demographic and Health Surveys data, for which the ethical responsibility is entirely of the institutions that conducted the surveys in each country, eliminating the requirement of this study's ethical approval.

