## Supplementary material for "Association between women’s empowerment and demand for family planning satisfied among Christians and Muslims in multi-religious African countries"

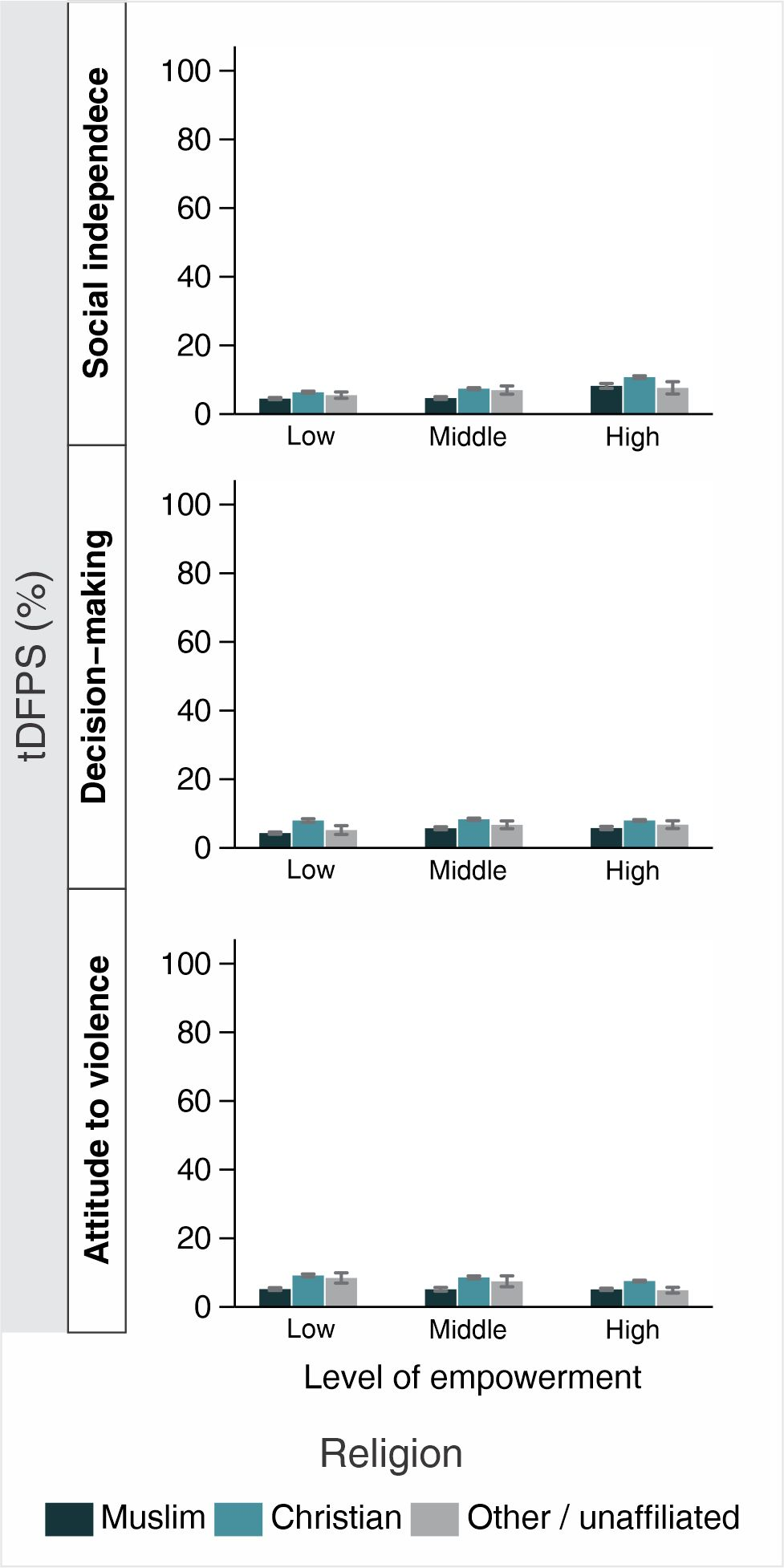

Supplementary Figure 1. Demand for family planning satisfied by traditional methods according to religion and women’s empowerment.

Supplementary Table 1. Demand for family planning satisfied according to religion, type of method, and women’s empowerment in the selected countries.

|  | Muslim | | Christian | | Interaction p value |
| --- | --- | --- | --- | --- | --- |
|  | IRR | 95% CI | IRR | 95% CI |  |
|  | **Traditional methods** | | | |  |
| Social independence |  |  |  |  | 0.000 |
| Low | 1.00 |  | 1.00 |  |  |
| Middle | 1.12 | (0.91; 1.38) | 1.25 | (1.04; 1.50) |  |
| High | 1.68 | (1.28; 2.21) | 2.11 | (1.78; 2.49) |  |
| Attitude to violence |  |  |  |  | 0.000 |
| Low | 1.00 |  | 1.00 |  |  |
| Middle | 0.88 | (0.55; 1.40) | 1.09 | (0.91; 1.30) |  |
| High | 0.95 | (0.56; 1.59) | 1.34 | (0.92; 1.93) |  |
| Decision-making |  |  |  |  | 0.000 |
| Low | 1.00 |  | 1.00 |  |  |
| Middle | 1.43 | (1.13; 1.81) | 1.28 | (0.98; 1.67) |  |
| High | 1.87 | (1.47; 2.36) | 1.52 | (1.20; 1.93) |  |
|  | **Modern methods** | | | |  |
| Social independence |  |  |  |  | 0.000 |
| Low | 1.00 |  | 1.00 |  |  |
| Middle | 1.13 | (0.95; 1.34) | 1.02 | (1.00; 1.04) |  |
| High | 1.38 | (1.02; 1.85) | 1.02 | (0.96; 1.08) |  |
| Attitude to violence |  |  |  |  | 0.000 |
| Low | 1.00 |  | 1.00 |  |  |
| Middle | 1.20 | (1.09; 1.33) | 1.03 | (0.98; 1.08) |  |
| High | 1.29 | (1.12; 1.49) | 1.06 | (1.01; 1.11) |  |
| Decision-making |  |  |  |  | 0.000 |
| Low | 1.00 |  | 1.00 |  |  |
| Middle | 1.27 | (1.14; 1.43) | 1.10 | (1.04; 1.16) |  |
| High | 1.45 | (1.22; 1.73) | 1.13 | (1.05; 1.22) |  |
